# Wellbeing Ways: Protocol for Assessing and Supporting Aboriginal and Torres Strait Islander Students’ Wellbeing in Queensland Secondary Schools

**DOI:** 10.64898/2026.01.23.26344683

**Authors:** Kate Anderson, Neelam Malik, Darren Garvey, Kirsten Howard, Michelle Dickson, Gail Garvey

## Abstract

**Background:** Aboriginal and Torres Strait Islander youth belong to culturally rich and resilient communities, yet they continue to experience significant health and wellbeing disparities. Despite these challenges, there is a critical gap in culturally appropriate tools to assess and support the wellbeing of Aboriginal and Torres Strait Islander youth in school settings. The Wellbeing Ways project addresses this gap by working with secondary schools to implement What Matters 2Youth (WM2Y), a newly developed, strengths-based wellbeing measure grounded in the perspectives of Aboriginal and Torres Strait Islander youth (aged 12-17 years), and co-designing place-based supports.

**Methods:** Wellbeing Ways is a three-year project being conducted in partnership with four diverse Queensland secondary schools. The project employs a four-phase mixed methods design guided by co-design principles and the i-PARiHS implementation framework. Phase 1 involves pre-implementation yarning workshops to co-design culturally safe implementation strategies; Phase 2 delivers longitudinal administration of WM2Y; Phase 3 conducts a post-implementation evaluation; and Phase 4 involves co-designing place-based wellbeing resources and referral pathways with students, staff, Elders, and service providers, informed by WM2Y data and community priorities.

**Ethics and Dissemination:** Ethics approvals have been obtained from relevant Human Research Ethics Committees. Findings will be disseminated through peer-reviewed publications, community presentations, and policy briefs, ensuring Aboriginal and Torres Strait Islander communities are primary audiences for all outputs.

## 1. Background

Aboriginal and Torres Strait Islander people make up 3.8% of the total Australian population, with a total estimated population of 983,700.[1] A notable feature of the Aboriginal and Torres Strait Islander population is its youth: one-third (33.1%) are under the age of 15, and the median age is just 24 years— significantly younger than the national median.[1] Aboriginal and Torres Strait Islander youth belong to culturally rich and resilient communities, underpinned by thousands of years of continuous cultural practice and knowledge. Their wellbeing is deeply embedded in cultural identity, community connections, and spiritual relationships with Country.[2] This culturally grounded concept of wellbeing encompasses physical, emotional, spiritual, cultural, and relational dimensions of health, in contrast to biomedical models that emphasise individual pathology.[2]

Despite the cultural strengths and resilience of Aboriginal and Torres Strait Islander communities, Aboriginal and Torres Strait Islander youth continue to face profound disparities across health, education, and justice systems.[3] Rates of psychological distress among Aboriginal and Torres Strait Islander youth are nearly twice as high as their non-Aboriginal and Torres Strait Islander peers.[4, 5] Suicide stands as the leading cause of death for Aboriginal and Torres Strait Islander people aged 15-24,[6] with rates significantly higher than for non-Aboriginal and Torres Strait Islander youth.[4, 6] Educational disadvantage is also a pressing issue, with Year 12 attainment for Aboriginal and Torres Strait Islander youth at 66% in 2021 compared to 96% for non-Aboriginal and Torres Strait Islander youth.[7]

### 1.1 The Need for Culturally Grounded Wellbeing Measurement

Accurate, culturally appropriate data is essential to inform and evaluate programs and policies aimed at improving the wellbeing of Aboriginal and Torres Strait Islander youth.[5, 8] However, a critical barrier to progress is the lack of culturally appropriate measures to assess wellbeing. Current wellbeing measures are predominantly based on Western biomedical constructs and do not capture the holistic, relational understandings of wellbeing held by Aboriginal and Torres Strait Islander communities.[8]

The 2020 National Agreement on Closing the Gap emphasises that progress must be Aboriginal and Torres Strait Islander-led, with cultural views and values prioritised.[9] This requires a substantial shift in how the wellbeing of Aboriginal and Torres Strait Islander youth is understood, measured, and supported. The most effective wellbeing measures are those that have been co-designed with communities, reflect cultural identity and lived experience, and are validated within Aboriginal and Torres Strait Islander contexts.[10]

### 1.2 The Role of Schools in Supporting Wellbeing

Schools are uniquely positioned to play a critical role in fostering mental health, social and emotional wellbeing, and early intervention for Aboriginal and Torres Strait Islander students.[11] Culturally safe, inclusive school settings that affirm identity and support cultural expression are central to promoting positive outcomes.[9] However, the ability of schools to deliver effective support is closely tied to student engagement, and many Aboriginal and Torres Strait Islander students face barriers that disrupt their connection to school, including experiences of racism, cultural alienation, and systemic marginalisation.[12, 13]

The relationship between school engagement and wellbeing is complex and reciprocal. While education has the potential to enhance mental health and resilience, the ability of youth to attend and thrive at school is often dependent on their wellbeing and that of their families and communities.[14] Without valid, culturally responsive measures of student wellbeing, it is difficult to assess the effectiveness of school-based interventions or tailor supports to meet specific needs.[10]

### 1.3 The What Matters 2Youth Measure

To address these gaps in wellbeing measurement, the What Matters 2Youth (WM2Y) measure was developed and validated through extensive co-design with Aboriginal and Torres Strait Islander youth across Queensland.[15] WM2Y is Australia’s first strengths-based, culturally grounded wellbeing measure for Aboriginal and Torres Strait Islander youth aged 12-17 years. The measure comprises items across eight domains: Belonging; Connection; Care and Self-care; Safe spaces; Aspirations; Independence; Health; and Respect and Racism.[16]

The Wellbeing Ways project represents a critical step in translating this wellbeing research into practice. By implementing WM2Y in Queensland secondary schools and co-designing place-based supports informed by student wellbeing data, this project aims to develop sustainable and scalable systems of wellbeing measurement and culturally responsive support for Aboriginal and Torres Strait Islander youth.

## 2. Methods

### 2.1 Research Team

As a research team, we are conscious of the importance of our backgrounds and perspectives, and reflexively consider how such factors may influence our approach to research.[17, 18] Our author team comprised Aboriginal and Torres Strait Islander Peoples (DG, MD, GG) and non-Indigenous researchers (KA, NM, KH).

### 2.2 Aboriginal and Torres Strait Islander Governance

Aboriginal and Torres Strait Islander leadership and governance throughout the project is a foundational element of Indigenist methodologies and equitable processes. Aboriginal and Torres Strait Islander governance has been established at each of the participating school sites, ensuring that decision-making, cultural oversight, and project direction are guided by Aboriginal and Torres Strait Islander voices and perspectives. Governance structures include school principals, Community Education Counsellors, regional program managers and coordinators, local government representatives, community Elders, and Aboriginal and Torres Strait Islander education staff. These governance arrangements ensure that the project is accountable to Aboriginal and Torres Strait Islander communities at each site, that cultural protocols are respected, and that implementation approaches are responsive to local contexts and priorities. Aboriginal and Torres Strait Islander staff and community representatives participate in all phases of the project, from co-designing implementation strategies to interpreting findings and guiding dissemination. This governance model embodies the principle that research with Aboriginal and Torres Strait Islander communities must be led by and accountable to those communities, ensuring that the Wellbeing Ways project generates knowledge and outcomes that are meaningful, culturally safe, and beneficial to Aboriginal and Torres Strait Islander students and families.

### 2.3 Indigenist Methodology

The current study is described in detail below as per the requirements of the *Consolidated criteria for strengthening reporting of health research involving Indigenous peoples* (the CONSIDER statement).[19] The study will employ a strengths-based, Indigenist methodology [20, 21] to understand how best to support the wellbeing of Aboriginal and Torres Strait Islander youth. Given the enduring process of colonial disempowerment and marginalisation of Aboriginal and Torres Strait Islander Peoples in Australia, the use of Indigenist methodologies and a strengths-based approach was a purposeful decision to avoid further subjugation of this group. We will use an Indigenist research methodology to ensure that Aboriginal and Torres Strait Islander youth’s voices guide all aspects of the research. Our research team has extensive experience in using Indigenist research methods to ensure that our work contributes to improving the outcomes and experiences of Aboriginal and Torres Strait Islander Peoples. Our approach will be guided by the following principles: ensuring Aboriginal and Torres Strait Islander voices and perspectives are prioritised and privileged throughout all aspects of the research process and our work; building Aboriginal and Torres Strait Islander Peoples’ research capacity and developing future research leaders; and facilitating collaboration through engaging and connecting with a range of key stakeholders.

### 2.4 Study design

The Wellbeing Ways project employs a three-phase mixed methods design, following principles of co-design with Aboriginal and Torres Strait Islander communities and guided by the i-PARiHS (integrated-Promoting Action on Research Implementation in Health Services) framework.[22] The i-PARiHS framework emphasises the interaction between evidence, context, and facilitation in successful implementation, making it well-suited for this community-engaged research.[23]

The project is being conducted in partnership with four Queensland secondary schools that serve significant populations of Aboriginal and Torres Strait Islander students. Co-design principles guide all phases of the work, ensuring that Aboriginal and Torres Strait Islander students, families, educators, and community members are central to decision-making and knowledge generation. For timeline of the phases refer to Table 1.

**Table 1:** Timeline of project phases.

|  | 2024 |  |  |  | 2025 |  |  |  | 2026 |  |  |  | 2027 |  |  |  |
| --- | --- | --- | --- | --- | --- | --- | --- | --- | --- | --- | --- | --- | --- | --- | --- | --- |
|  | Q1 | Q2 | Q3 | Q4 | Q1 | Q2 | Q3 | Q4 | Q1 | Q2 | Q3 | Q4 | Q1 | Q2 | Q3 | Q4 |
| HREC DOE approvals |  |  | X |  |  |  |  |  |  |  |  |  |  |  |  |  |
| Recruitment |  |  |  | X | X | X | X | X |  |  |  |  |  |  |  |  |
| **Phase 1:** Pre-implementation workshops |  |  |  |  |  | X | X | X |  |  |  |  |  |  |  |  |
| **Phase 2:** WM2Y implementation |  |  |  |  |  |  |  |  | X | X | X | X |  |  |  |  |
| **Phase 3:** Post implementation evaluation workshops |  |  |  |  |  |  |  |  |  |  |  |  | X |  |  |  |
| **Phase 4:** Co design workshops and resource/ program development and implementation |  |  |  |  |  |  |  |  |  |  |  |  |  | X |  |  |

The four phases of the project are:

Phase 1: Pre-implementation yarning workshops with school staff to co-design culturally safe implementation strategies suitable for each school setting

Phase 2: Longitudinal administration of WM2Y in four Queensland secondary schools over 12 months

Phase 3: Post-implementation evaluation of Phase 2 with school staff

Phase 4: Co-designing place-based wellbeing resources and referral pathways with students, staff, Elders, and service providers, informed by WM2Y data and community priorities

#### 2.4.1 Phase 1: Pre-Implementation Yarning Workshops

Prior to implementation, the research team will facilitate yarning workshops with school staff, Aboriginal and Torres Strait Islander education workers, and community stakeholders at each school site. These workshops will co-design locally relevant implementation strategies that respond to the unique context, resources, and cultural protocols of each school. Topics will include:

- Appropriate timing, frequency, and settings for WM2Y administration
- Culturally safe approaches to student engagement and consent
- Staff training needs and capacity-building
- Data management, confidentiality, and ethical protocols

#### 2.4.2 Phase 2: Longitudinal WM2Y Administration

Following co-design of implementation plans, WM2Y will be administered longitudinally over a 12-month period at each participating school. Administration will be led by trained school staff with support from the research team. The research team will monitor implementation fidelity, acceptability, and cultural appropriateness throughout this phase, adjusting as needed based on ongoing feedback from school communities.

#### 2.4.3 Phase 3: Post-Implementation Evaluation

Following the 12-month implementation period, post-implementation yarning workshops will be conducted with school staff and students to evaluate the process and outcomes. These workshops will explore:

- Acceptability and cultural safety of WM2Y administration
- Utility of wellbeing data for informing school-based supports
- Barriers and enablers to sustainable implementation
- Recommendations for refinement and scale-up

Quantitative WM2Y data will be analysed to identify patterns in student wellbeing, changes over time, and domains of particular strength or concern. These findings will inform the co-design work in Phase 4.

#### 2.4.4 Phase 4: Co-design of Place-Based Wellbeing Supports

Phase 4 involves the co-design of culturally responsive, place-based wellbeing resources, activities, and referral pathways tailored to each school context.[24] This phase will be informed by WM2Y data, community priorities, and the strengths and needs identified through yarning workshops.

Co-design workshops will engage Aboriginal and Torres Strait Islander students, families, Elders, educators, and local service providers in identifying local strengths, wellbeing priorities, and gaps in support. Participants will collaboratively develop and pilot wellbeing activities, resources, and referral pathways that are meaningful, culturally safe, and sustainable.

The feasibility, cultural safety, and effectiveness of co-designed supports will be evaluated through ongoing feedback from participants, ensuring that approaches are responsive to student and community needs.

### 2.5 Study Setting and Participants

The Wellbeing Ways project is being conducted in partnership with four Queensland secondary schools serving significant populations of Aboriginal and Torres Strait Islander students. Schools were selected based on their commitment to Aboriginal and Torres Strait Islander student wellbeing, existing relationships with the research team, and interest in implementing routine wellbeing assessment.

Participants include:

- Aboriginal and Torres Strait Islander students aged 12-17 years enrolled at participating schools
- School staff including principals, wellbeing coordinators, Aboriginal and Torres Strait Islander education workers, and classroom teachers
- Aboriginal and Torres Strait Islander families and community Elders
- Local service providers supporting youth wellbeing

### 2.6 Data Collection and Analysis

The project employs mixed methods data collection including:

- Quantitative WM2Y wellbeing data collected longitudinally
- Qualitative data from yarning workshops and co-design sessions, including audio recordings, field notes, and visual materials
- Implementation process data including attendance records, fidelity checklists, and researcher observations

Quantitative data will be analysed using descriptive statistics and, where appropriate, longitudinal modelling to examine changes in wellbeing over time. Qualitative data will be analysed thematically, with coding and interpretation conducted in partnership with Aboriginal and Torres Strait Islander research team members and community representatives to ensure cultural integrity and accuracy.

### 2.7 Ethics and Dissemination

The research methodology and conduct adhered to the ethical principles outlined in the AIATSIS Code of Ethics for Aboriginal and Torres Strait Islander Research,[25] ensuring respectful and culturally appropriate engagement with Indigenous knowledge and communities. Ethics approvals were obtained for this phase of the study from: The University of Queensland Human Research Ethics Committee (ref: 2024/HE000020); Far North Queensland Human Research Ethics Committee (re: HREC/2024/QCH/105828); and Queensland Department of Education (ref: 550/27/2834). Each team member attending sites attained the relevant jurisdictions’ Working with Children Check.

All participants provided written informed consent. For youth participants, written consent was obtained from a parent or legal guardian, and youth assent was obtained using a written assent form. Data are stored securely in accordance with institutional requirements, and strict confidentiality protocols protect participant privacy. Community governance structures guide all aspects of the research, including data ownership, interpretation, and dissemination.

Findings will be disseminated through multiple channels to ensure impact across research, policy, and practice domains:

- Peer-reviewed journal publications;
- Community presentations and feedback sessions at each participating school;
- Policy briefs for Queensland education and health sectors;
- Conference presentations at national and international forums; and
- Plain language summaries and resources developed in partnership with student participants.

Aboriginal and Torres Strait Islander communities are positioned as primary audiences for all research outputs, with dissemination approaches prioritising accessibility, cultural appropriateness, and practical utility for schools and families.

2.8 Timeline of Study

### 2.8 Expected Outcomes and Impact

The Wellbeing Ways project is expected to deliver significant outcomes for Aboriginal and Torres Strait Islander youth wellbeing, school practice, and research translation:

- Demonstrated feasibility and acceptability of culturally grounded wellbeing measurement in Queensland secondary schools;
- Real-time wellbeing data to inform targeted, responsive support for Aboriginal and Torres Strait Islander students;
- Co-designed, place-based resources and referral pathways that reflect community priorities and cultural values;
- Enhanced capacity of school staff to assess and support Aboriginal and Torres Strait Islander student wellbeing;
- Evidence base for scaling WM2Y implementation across other Queensland schools; and
- Contribution to Closing the Gap targets through improved understanding and support of Aboriginal and Torres Strait Islander youth wellbeing.

By centring Aboriginal and Torres Strait Islander voices, perspectives, and leadership throughout, this project aims to shift how wellbeing is understood and supported in school settings, creating more equitable, culturally responsive systems of care for Aboriginal and Torres Strait Islander youth.

## 3. Conclusion

The Wellbeing Ways project represents a critical advancement in Aboriginal and Torres Strait Islander youth wellbeing research and practice. By implementing the first culturally grounded wellbeing measure for Aboriginal and Torres Strait Islander youth in Queensland secondary schools, and co-designing place-based supports informed by student data and community priorities, this work addresses longstanding gaps in how wellbeing is understood, measured, and supported.

Guided by co-design principles and grounded in Aboriginal and Torres Strait Islander definitions of wellbeing,[24] this project offers a pathway toward more equitable, responsive, and sustainable approaches to supporting the social and emotional wellbeing of Aboriginal and Torres Strait Islander youth. The outcomes will inform policy and practice across Queensland and contribute to national efforts to close gaps in health, education, and wellbeing outcomes for Aboriginal and Torres Strait Islander youth.

## Data Availability

Data cannot be shared publicly because of participant privacy concerns.

## Author Contributions

KA, study conception, study design, manuscript drafting, review and revision. NM, study design, review and revision. DG, study design, manuscript review and revision. KH, study conception, study design, and manuscript review and revision. MD, study design, manuscript review and manuscript revision. GG, study conception, study design, manuscript review and revision. All authors have read and approved the final manuscript.

